# Investigating inequalities in HIV testing in sub-Saharan Africa: insights from a spatial analysis of 25 countries

**DOI:** 10.1101/2022.10.20.22281320

**Authors:** Pearl Anne Ante-Testard, Gabriel Carrasco-Escobar, Tarik Benmarhnia, Laura Temime, Kévin Jean

**Author notes:** Corresponding author: Pearl Anne Ante-Testard, Laboratoire MESuRS, Conservatoire national des Arts et Métiers, 292 rue Saint Martin, 75003 Paris, France, or. E-mail addresses of authors: P.A.A.-T. or G.C.-E. T.B. L.T. K.J.

## Abstract

**Introduction:** We aim to explore spatial variations in socioeconomic inequalities in self-reported recent HIV testing uptake in sub-Saharan Africa (SSA) at different geographical scales, in order to identify potential geographical hotspots of inequalities. Additionally, to evaluate the potential benefits of HIV testing programs, we assess whether local levels of HIV testing match the local levels of HIV prevalence.

**Methods:** We analyzed data from 25 countries in SSA with Demographic and Health Surveys between 2011 and 2019. We quantified socioeconomic inequalities in self-reported HIV testing in the last 12 months with both the Slope Index of Inequality (SII) and Relative Index of Inequality (RII) in different geographical scales to capture sex-specific within-country spatial variations. We also conducted sampling cluster-level analyses based on the Local Indicator of Spatial Association to consider the autocorrelation in SII and RII across SSA countries. To assess the spatial efficiency of HIV testing programs, we measured the correlation between recent HIV testing uptake and HIV prevalence through Pearson correlation across geographical scales.

**Results:** We observed pro-rich inequalities on both absolute and relative scales in recent HIV testing in the majority of SSA countries in female and male participants at the national level. We also identified inequalities at subnational levels. Within- and between-country heterogeneities in sex-specific inequalities on both inequality scales and their respective spatial distributions varied depending on the scale used. Clustering of high absolute and relative inequalities were mostly observed in Western and Central Africa with a few regions in Eastern and Southern Africa. Despite significant sex-specific correlations between HIV testing and HIV prevalence in all countries when assessed at the national level, we report an absence of such a correlation at fine scale in 39 of 50 sex-country combinations.

**Conclusions:** These findings highlight the importance of investigating the spatial variability of various HIV indicators and related inequalities. Results may help local, national and international policymakers to prioritize areas and socioeconomic subgroups in need of HIV testing services. Our results also show the need to monitor efficiency of HIV testing programs in relation to HIV risk at subnational levels as a complementary to national estimates.

## Introduction

The role of HIV testing in the fight against HIV/AIDS is crucial since it is the gateway to HIV prevention and care, especially in sub-Saharan Africa (SSA), the global epicenter of the disease. Over the years, testing has scaled up due to the increasing availability of antiretroviral therapy (ART) and in response to the UNAIDS 90-90-90 and 95-95-95 targets by 2020 and 2030, respectively [1,2].

Socioeconomic inequalities have been reported in HIV testing uptake in SSA. Numerous studies have found that people in higher socioeconomic position (SEP) were more likely to seek HIV testing or know their HIV status [3–8]. However, most of these studies assessed testing inequalities at the national level. Very few studies have analyzed the spatial distribution of these inequalities. To the best of our knowledge, such a local analysis has only been performed in a single country, Malawi [9].

Spatial analysis at local scales has proved useful for the control and prevention of infectious diseases, by uncovering spatial variations in the transmission or the access to prevention [10–12]. Regarding HIV, it has helped identify high transmission areas [13] and understand access difficulties to healthcare facilities in underserved areas in Africa [14]. However, it has not been frequently utilized in monitoring inequalities in the HIV response, particularly in HIV testing. Notably, uptake of HIV testing has been reported to be higher in countries with the greatest HIV burden at the national level. Nevertheless, whether this association also exists at a finer scale has not been empirically assessed. Observing such spatial variations is important not only for ensuring equity in epidemic control but also for prioritizing areas with the greatest burden in terms of infection and/or inequalities [15]. Mapping HIV testing uptake and their inequalities across different geographical scales and identifying their local hotspots has become especially relevant in a context of decreasing international funding for the HIV response.

Here, we aim: i) to explore spatial variations in socioeconomic inequalities in HIV testing uptake across geographical scales; ii) to identify geographical hotspots for such inequalities in several SSA countries; and iii) to assess spatial correlations between HIV testing and HIV prevalence at the national and subnational levels as a way to approximate the potential benefits of existing programs.

## Methods

### Study design and data sources

We carried out a multi-country analysis of cross-sectional surveys in SSA countries, namely the Demographic and Health Surveys (DHS). The DHS are nationally representative surveys regularly conducted in the Global South collecting information on a broad range of indicators such as sociodemographic indicators, maternal and child health, malaria and HIV/AIDS. They are based on a two-stage sampling design, with Primary Sampling Units (PSU) selected in the first stage and households in the second. Women aged 15-49 years and men aged 15-54/59 years in participating households are eligible. Depending on the survey, data for men for the HIV indicators, biomarkers or both might may be collected only from a sub-sample of the selected households. Some DHS include HIV serological surveys in which participants are asked for consent to be tested for HIV, which is done anonymously in most of the surveys. Individuals who consented are interviewed face-to-face by trained interviewers who use a standard questionnaire. The agencies and research institutes that conducted the surveys were responsible for acquiring ethical clearance in each country.

DHS Global Positioning System coordinates were obtained from the DHS database. These coordinates were intentionally and randomly displaced to ensure confidentiality of the respondents. Urban clusters were displaced between 0 and 2 kilometers (km), while rural clusters were displaced between 0 and 5 km. The spatial boundaries were also obtained from the DHS spatial database repository (https://spatialdata.dhsprogram.com/home/). The DHS data were linked to these spatial data.

For our analysis, we selected SSA countries with available DHS surveys between 2011 and 2019 that contained the variables of interest and spatial data. We selected the more recent survey (as of November 2021) in countries with more than one eligible survey.

### Data/ Variables

SEP was defined as the relative rank of the participants in the cumulative distribution of the DHS wealth index. The wealth index is a composite measure of living standards based on household assets and living characteristics. The DHS divided the wealth index homogenously into quintiles from poorest to richest.

The outcome of interest was the self-report of recent (< 12 months) HIV testing uptake. Participants were asked if they recently had an HIV test and the time since last test. Being HIV positive was defined as testing positive in the serological survey.

### Statistical analysis

First, for each country and sex, we calculated the HIV prevalence and the proportion of recent HIV testing uptake while accounting for survey design and sampling weights at different geographical scales: i) national, ii) first administrative subnational level (hereafter, province), and iii) PSU level (hereafter referred to as “fine scale”).

Second, we measured national-, province-, and PSU-level socioeconomic inequalities both on the absolute and relative scales. We estimated the Slope Index of Inequality (SII) and the Relative Index of Inequality (RII) to assess the absolute and relative inequalities, respectively [16]. It is highly recommended to report inequalities on both scales as conclusions may diverge depending on the scale used [17].

At the national and province levels, both indicators were obtained by fitting a modified Poisson regression (with robust variance) with a log link function [18] to estimate the association between recent HIV testing at each wealth level and the hierarchical ranking of wealth. Generalized estimating equations were used to account for the clustering of observations [19]. The SII represents the absolute difference in the predicted proportions between the richest and the poorest participants, whereas the RII expresses the ratio of the predicted outcomes between these two extremes.

Due to the smaller sample sizes at the PSU-level (at least 10 individuals), we fitted a linear regression to estimate fine-scale inequality indicators. When performed on small samples, regressions may lead to negative RII values, which we truncated to zero when estimating the fine-scale RII.

Third, spatial autocorrelations of the fine-scale SII and RII across SSA and between sexes were assessed using the local *Getis-Ord Gi\** statistic for PSUs with a sample size of at least 10. The Getis-Ord Gi* statistic identifies local patterns and clusters of high- or low-inequality across countries that may not be evident when using global statistics [20]. More specifically, a distance-based neighborhood structure was used for its computation. Neighboring PSUs were defined based on the distance *d* that assigns a number of nearest neighbors (*k*) to each PSU. We selected the number of nearest neighbors that gave high spatial autocorrelation based on a global Moran’s I statistic for each sex and inequality indicator. We categorized the Gi*\** statistic based on the sign (cold- or hotspot for negative and positive signs, respectively) and percentile (90%, 95%, 99%) to avoid bias due to multiple and dependent tests [21].

We assessed the spatial correlation of the proportion of recent HIV testing and level of HIV prevalence across various geographical scales through Pearson correlation. Indeed, HIV prevalence drives, at least partly, the local risk of incident HIV infection as it reflects the probability for one’s sexual partner to be infected by HIV [22].

### Sensitivity analysis

SII and RII were calculated for PSUs with a sample size of at least 20 and 30 and their local spatial autocorrelations were assessed as sensitivity analysis. We also conducted similar analyses for countries with surveys between 2011 and 2014 and for countries with surveys between 2015 and 2019 to assess possible temporal trends in the spatial distribution of inequalities.

## Results

### Study population characteristics

Twenty-five countries were eligible based on data availability between 2011 and 2019: Angola, Burundi, Cameroon, Chad, Côte d’Ivoire, Democratic Republic of Congo (Congo DR), Ethiopia, Gabon, Ghana, Guinea, Lesotho, Liberia, Malawi, Mali, Mozambique, Namibia, Rwanda, Senegal, Sierra Leone, Tanzania, Togo, Uganda, South Africa, Zambia, and Zimbabwe. Table 1 shows the summary statistics by country and sex. There was a total of 472,763 participants (311,652 women and 161,111 men) with 351,921 individuals (74.4%, 252,508 women and 99,413 men) from PSUs with a sample size of at least 10 and complete data (Table S1). The sample size in the provinces ranged between 275 and 11,342 among women and between 135 and 3, 236 among men. At a finer scale, sample size in PSUs ranged between 10 and 96 women and between 10 and 54 men. The distributions of PSU sample size and proportion of recent HIV testing are shown in Figure S1.

**Table 1.**
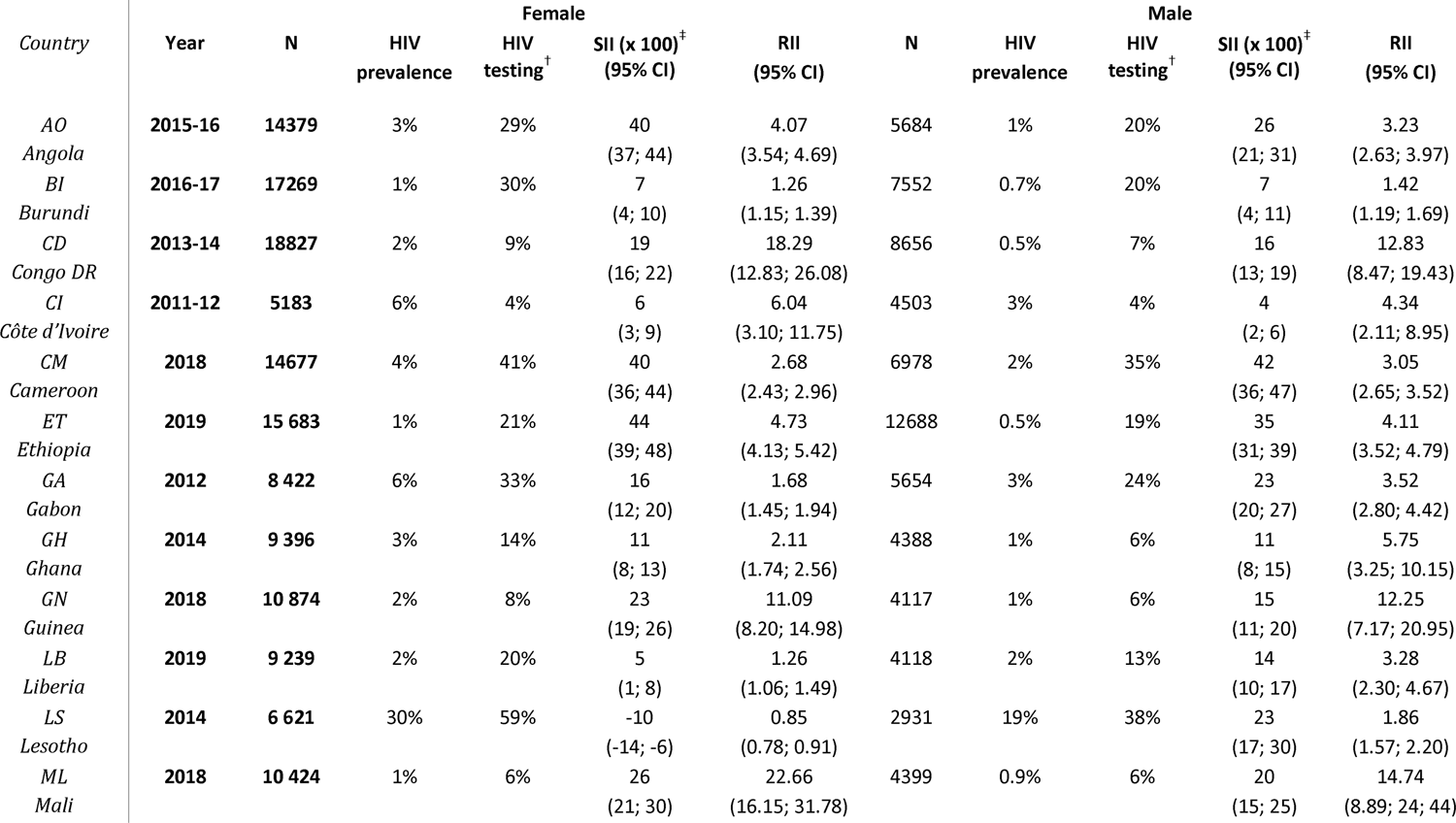
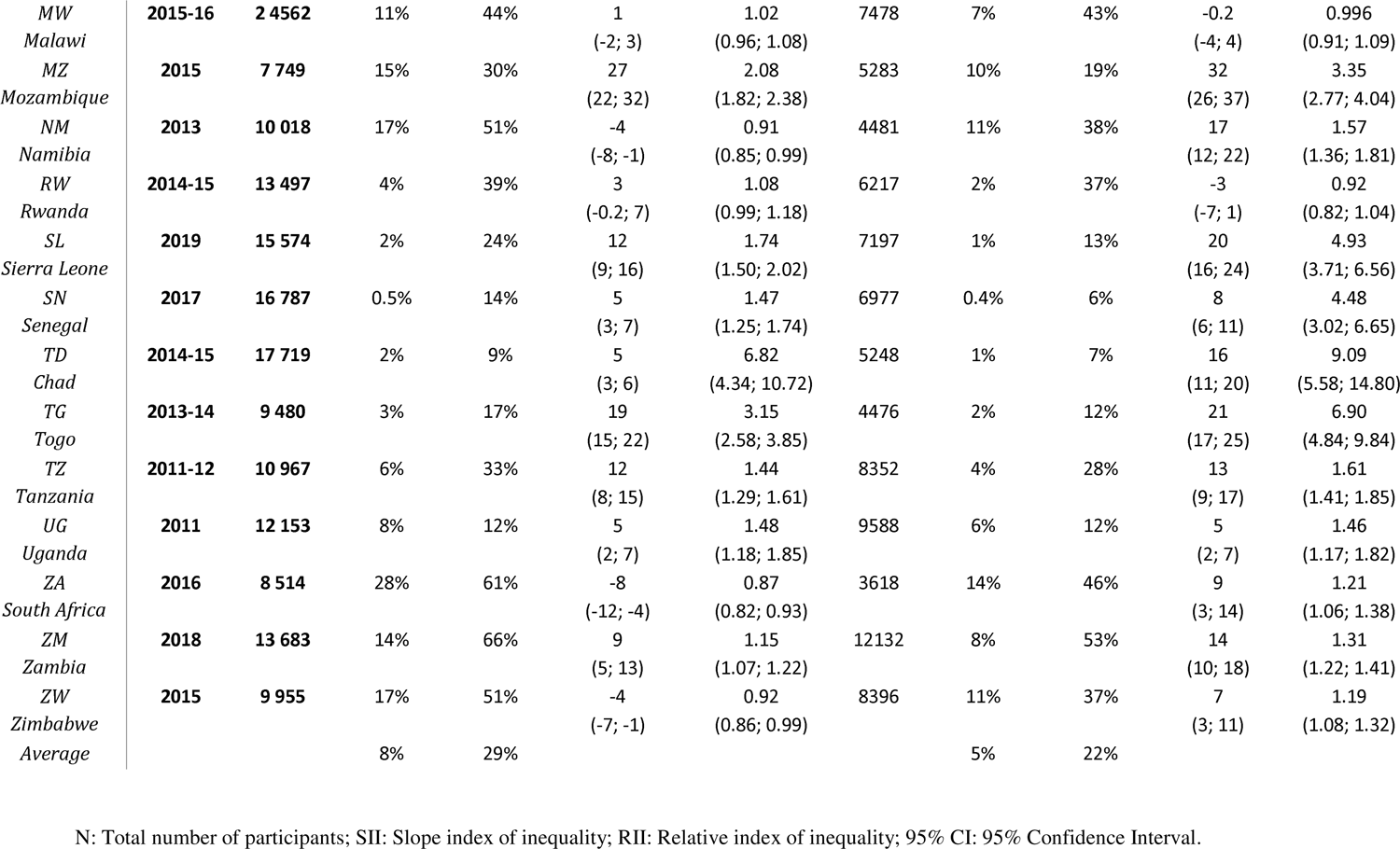
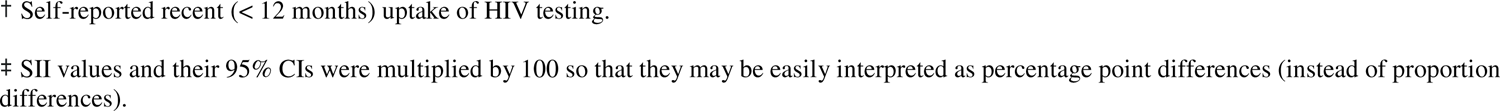
Summary estimates of national level HIV prevalence, HIV testing, absolute and relative inequalities in HIV testing uptake in the previous 12 months in 25 sub-Saharan African countries between 2011 and 2019 by country and sex.

### National-level estimates

Table 1 also presents the national estimates of HIV prevalence, testing, and inequalities in recent HIV testing. Overall, at the national level, HIV prevalence ranged from 0.5% (Senegal) to 30% (Lesotho) among women and from 0.4% (Senegal) to 19% (Lesotho) among men. Self-reported uptake of recent testing ranged from 4% (Côte d’Ivoire) to 66% (Zambia) among women and from 4% (Côte d’Ivoire) to 53% (Zambia) among men. Women tended to have higher HIV prevalence and proportion of recent testing than men.

On the absolute scale, we observed pro-rich absolute inequalities in recent testing in 19 of 25 countries for women and 23 of 25 countries for men (SII > 0) (Table 1). Absolute inequalities ranged between −8 (95% Confidence Interval [95% CI] −12; −4) percentage points (% points) in South Africa and 44 (39; 48) % points in Ethiopia among women. This means that the absolute difference between the richest and poorest quintiles was −8 (−12; −4) % points (i.e., pro-poor) among women in South Africa and 44 (39; 48) % points in Ethiopia (i.e., pro-rich). Meanwhile, among men, SII ranged between −3 (−7; 1) % points in Rwanda and 42 (0.36; 0.47) % points in Cameroon.

We observed pro-rich relative inequalities in 18 of 25 countries for women and 23 of 25 countries for men (RII > 1) (Table 1). Relative inequalities ranged between 0.85 (0.78; 0.91) in Lesotho and 22.66 (16.15; 31.78) in Mali among women. This translates to the richest women being 0.85 (0.78; 0.91) times as likely to report recent HIV testing than the poorest participants in Lesotho (i.e., pro-poor), while 22.66 times (16.15; 31.78) more likely in Mali (i.e., pro-rich). Among men, it ranged between 0.92 (0.82; 1.04) in Rwanda and 14.74 (8.89; 24.44) in Mali.

### Province-level estimates

The distribution of province-level HIV prevalence and proportion of recent HIV testing are mapped in Figure 1. We observed within- and between-country variations in their respective spatial distributions.

**Figure 1.**
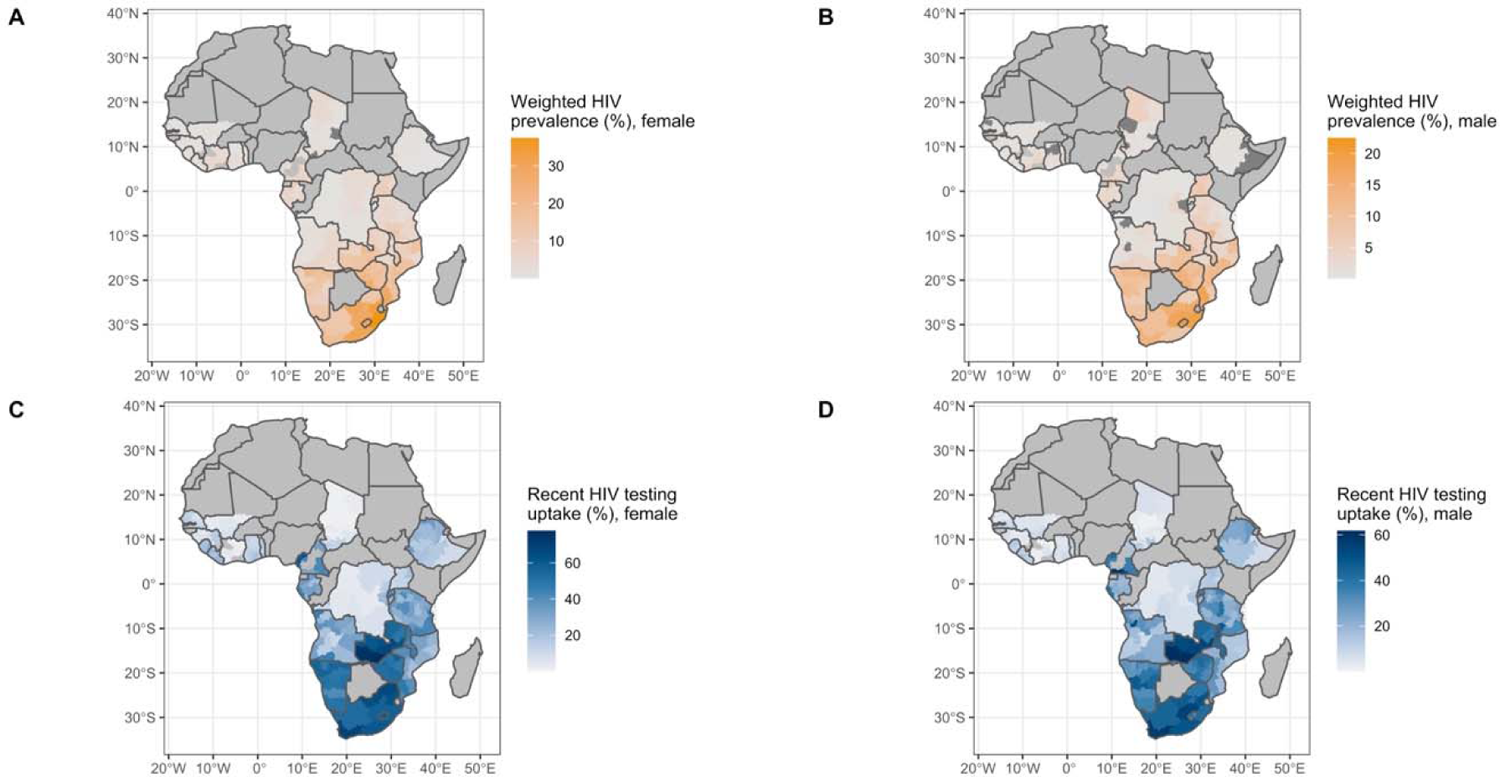
Province-level distribution of weighted HIV prevalence among A) female and B) male participants and weighted percentage of self-reported recent (<12 months) uptake of HIV testing among C) female and D) male participants. Dark grey colours indicate unavailability of the HIV biomarker. Missing polygons within the country indicate no data is available for this province from the Demographic and Health Surveys.

Figure 2 maps the provincial absolute and relative inequalities in recent HIV testing. We also observed spatial heterogeneities of these inequalities for all sexes and inequality scales. On the absolute scale, we observed pro-rich spatial distribution of SIIs in most of the provinces in SSA except for a few areas in Eastern and Southern Africa (ESA) such as South Africa, Namibia, and Malawi. On the relative scale, higher pro-rich relative inequalities were observed more frequently in Western and Central Africa (WCA), while lower inequalities tended to be observed in ESA.

**Figure 2.**
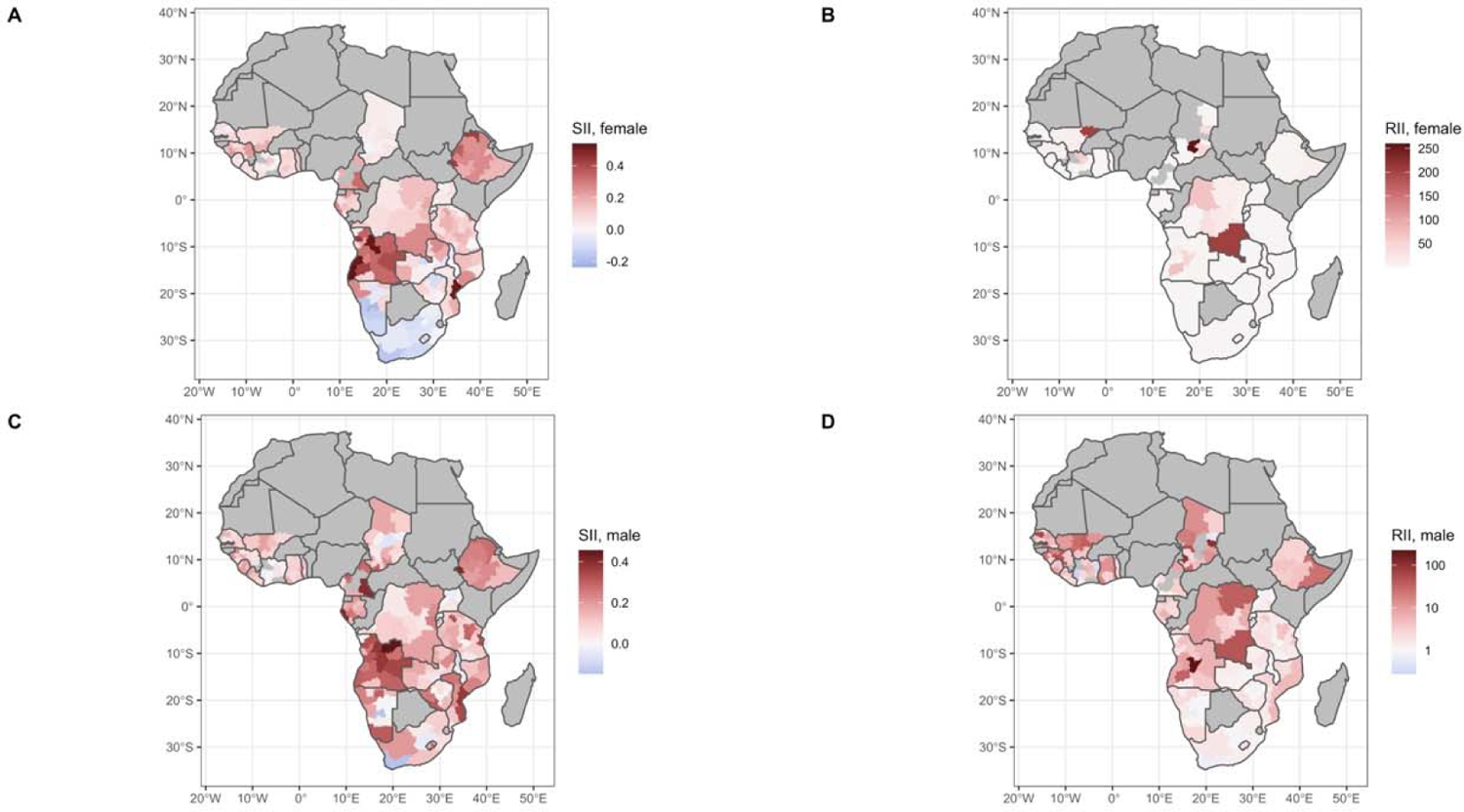
Province-level distribution of wealth-related inequalities in self-reported recent (<12 months) uptake of HIV testing at the A) and C) absolute scale and B) and D) relative scale among female and male participants, respectively, across 25 sub-Saharan African countries. SII, Slope Index of Inequality; RII, Relative Index of Inequality. Capped RII values between 0.1 and 300. Missing polygons in Chad represent provinces with extreme RII values (> 300 for females) and (< 0.1 for males).

### Spatial clustering analysis at fine scale

Global Moran’s I showed that using 1 or 2 nearest neighbors gave the highest spatial autocorrelation for both sexes and inequality indicators (Figure S2). For uniformity, we used *k=2* to calculate for the Getis-Ord Gi* statistics. Hotspots and coldspots of inequalities across SSA depended on the inequality scale used and sex (Figure 3). Overall, hotspots on both scales were more marked in WCA and a few ESA countries such as Ethiopia (for both sexes), Mozambique and Tanzania (only on relative scales for both sexes).

**Figure 3.**
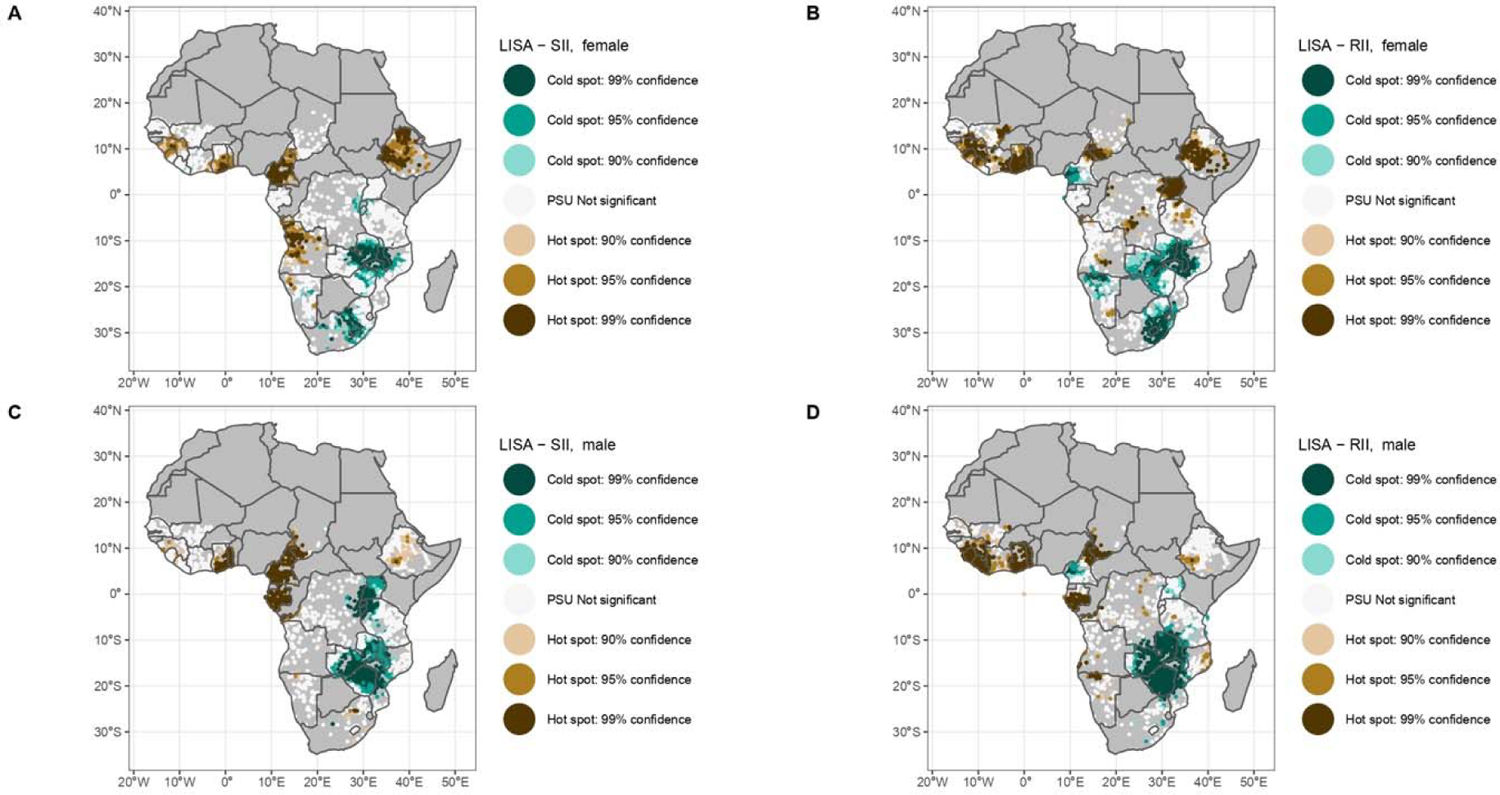
Local spatial autocorrelation of socioeconomic inequalities in self-reported recent (<12 months) uptake of HIV testing as Local Getis-Ord Gi* at Primary Sampling Unit (PSU) level (two nearest neighbors) across 25 sub-Saharan African countries between 2011 and 2019. Spatial clustering at the absolute scales among A) and C) absolute scales and B) and D) relative scales among female and male participants, respectively. Only PSUs with a sample size of at least 10 and more than one wealth quintile were included. LISA, Local Indicator of Spatial Autocorrelation; SII, Slope Index of Inequality; RII, Relative Index of Inequality.

On the absolute scale, we observed numerous pockets of high SII values in western Africa with a few areas in eastern Africa. Hotspots of absolute inequalities in recent HIV testing were observed in Cameroon, Ghana, Togo, and Ethiopia for both sexes. Coldspots or pockets of low SIIs were mostly observed in ESA including Zambia, Zimbabwe, Burundi, Rwanda, and small areas in Mozambique for both sexes and in South Africa for women.

On the relative scale, we observed hotspots of RII values mostly in WCA with a few areas in eastern Africa. Coldspots were noted in ESA including Burundi and Rwanda both sexes and in South Africa for women.

Moreover, we observed diverging patterns of hotspots and coldspots for each sex in the same country. There were hotspots of RIIs in Uganda among women, while coldspots in a few areas were noted among men. Contrastingly, areas in Namibia displayed coldspots among women, while hotspots were noted among men.

### Spatial correlation between HIV testing and HIV prevalence

We assessed the spatial correlation between HIV testing and HIV prevalence across geographical scales – whether testing services are reaching those with high HIV risk in the population. At the national level, HIV prevalence and proportion of recent HIV testing were found to be positively correlated among women and men (Figure S3).

However, this observation was not sustained at subnational levels. Within-country correlation of province-level HIV prevalence and proportion of testing showed that in many countries, both variables were uncorrelated (i.e., the level of HIV testing did not always match the magnitude of HIV prevalence). Out of 50 settings (2 sex-specific results for each of the 25 countries), we observed only 11 that had statistically significant positive correlations (p-value < 0.05) or what we termed as having “efficient HIV testing services” (Figure S4): both sexes (Côte d’Ivoire, Ethiopia, and Tanzania), females (Rwanda and Zambia) and males (Liberia, Lesotho, and Sierra Leone).

Similarly, Figure 4 shows that at the PSU level, HIV prevalence did not correlate with the level of recent HIV testing in many of the countries for both sexes. Only in Burundi (female), Mozambique (female), Namibia (male), Tanzania (both sexes), and Zambia (both sexes) did we observe a significant positive correlation. Meanwhile, in Lesotho, HIV prevalence and recent testing had a significant negative correlation (i.e., PSUs with higher HIV prevalence had lesser uptake of recent HIV testing).

**Figure 4.**
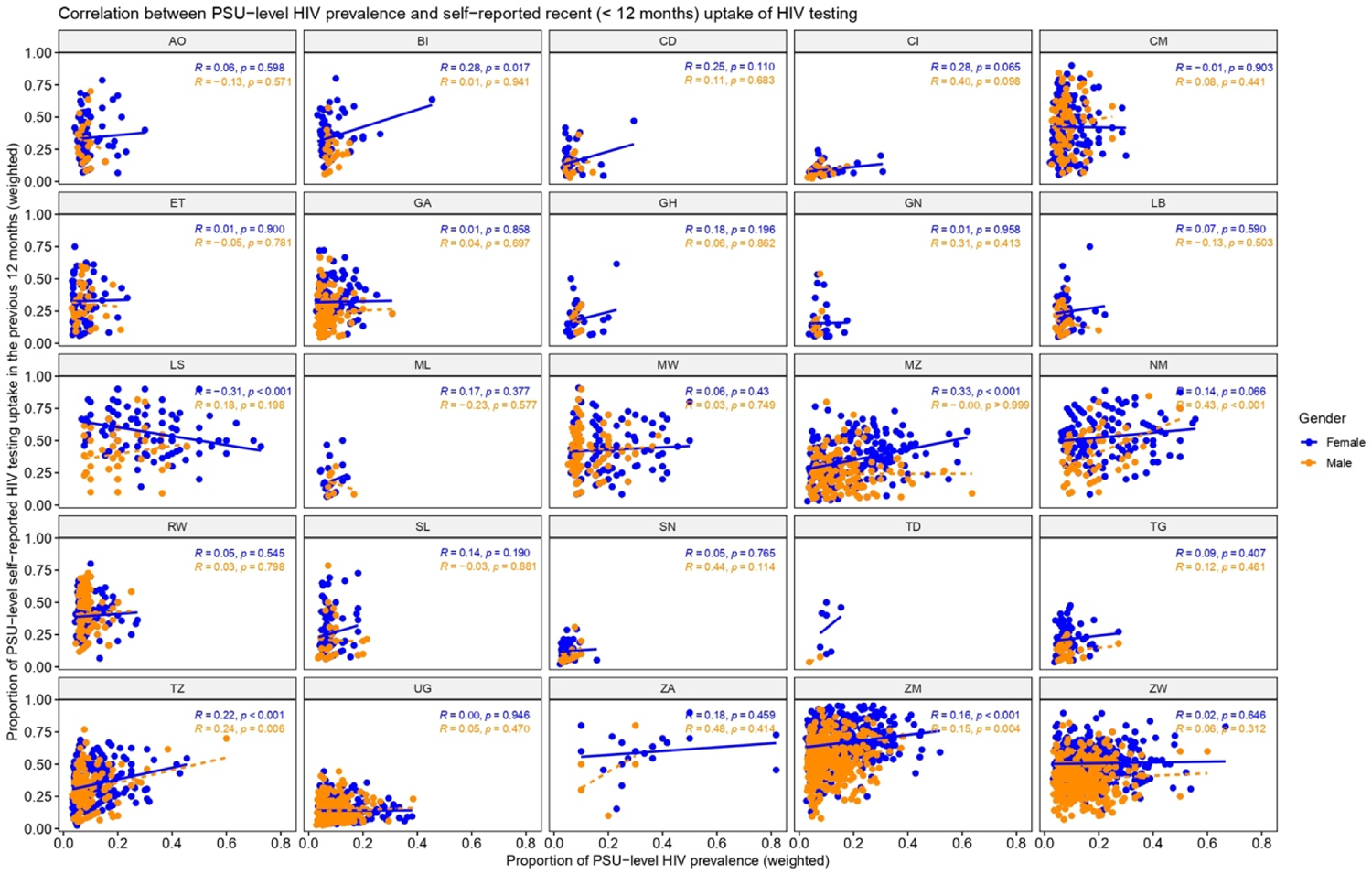
Correlation (Pearson correlation coefficient R and p-value) between the proportion of HIV prevalence and proportion of self-reported recent (< 12 months) uptake of HIV testing at the Primary Sampling Unit (PSU) level in 25 sub-Saharan African countries stratified by sex. Only included PSUs with a sample size equal or higher than 10 with both the HIV biomarker and HIV testing variables. There were not sufficient clusters to calculate the Pearson correlation R and p-values in Chad (TD). Each point represents a PSU. Refer to Table 1 for full country names.

### Sensitivity analysis

Results of the local spatial clustering analysis were consistent when sub-setting by cluster size of at least 20 or 30 (Figures S5 and S6). Areas with pockets of high and low inequalities were also consistent when conducting spatial clustering analysis across countries with surveys between 2011 and 2014 and between 2015 and 2019 separately (Figures S7 and S8).

## Discussion

In this study, we quantified and mapped absolute and relative socioeconomic inequalities of recent HIV testing at different geographical scales. We also conducted spatial clustering analysis of such inequalities and explored the spatial correlation between HIV testing and HIV prevalence at various geographical scales across SSA. Our results show existing inequalities at the national, province, and PSU levels. Heterogeneities in the spatial distribution of these inequalities at subnational levels and hotspot areas varied depending on the inequality scales and sex groups. Most hotspots of inequalities were observed in WCA with a few areas in ESA such as Ethiopia, Mozambique, and Tanzania. Meanwhile, coldspots were rather observed in ESA. We also revealed that, while HIV testing programs seemed efficient in reaching those with high risk of acquiring HIV at the national level, they seemed to be less efficient at the subnational levels in most of the countries. Indeed, the provinces and PSUs with higher recent testing uptake did not match the level of HIV prevalence in many of the countries for both sexes.

We tended to observe higher HIV prevalence and proportion of recent HIV testing among women and countries located in ESA. As expected, we report higher pro-rich inequalities among men and in WCA countries. These findings were consistent with previous studies [5,7]. Higher HIV burden among women may be explained by higher vulnerability than men in SSA due to several biological, societal, and economic factors [23].

We also highlighted that recent HIV testing was not shared equally across wealth levels in SSA (within and between countries) and that such inequalities were not randomly distributed across space. Indeed, our results also showed varying spatial patterns of recent HIV testing inequalities between absolute and relative scales and between women and men. This highlights the necessity for HIV testing programs to be tailored depending on the level of inequality to be addressed and the needs of each sex. We observed that in a few countries like Namibia, national-level inequality estimates showed low relative inequality for women and even pro-poor inequality on the absolute scale for both sexes suggesting that pro-rich inequalities are not a fatality and might be overcome. Nevertheless, hotspots of inequalities were still observed in some areas within these countries that achieved equitable HIV testing levels. This suggests that national-level inequality estimates may hide remaining pro-rich inequalities at lower scales.

However, one could argue that pro-rich inequalities are not necessarily unfair, especially if those with higher SEP also tend to be the ones who are more at-risk of acquiring HIV. Several early studies reported higher levels of HIV prevalence among higher SEP [24,25]. However, this initial inverse social gradient, an unusual feature as compared to most of the diseases, is likely in a process of reversion. Last-generation HIV prevalence surveys now report higher levels of HIV prevalence among the poorest in some countries such as Côte d’Ivoire, Ethiopia, Lesotho, and Zimbabwe. Moreover, a recent cohort study relying on a two-decade follow-up in rural Uganda documented a changing socioeconomic gradient over time, with a higher risk of incident HIV infection among the poorest [26]. Thus, suggesting that these pro-rich inequalities might indeed be unfair and contributing to the reversing social gradient that is probably ongoing in some settings.

There are various criteria for health programs that can be used to evaluate their public benefit which include efficiency. Efficiency is concerned with the optimal production and distribution of scarce health resources and is critical for sustainability and maximizing health gains [27]. In this study, we did not conduct a formal impact evaluation of HIV testing programs, but rather to investigate the spatial correlations between HIV testing and HIV prevalence across geographical scales to capture their potential benefits. Community-level HIV prevalence has been found to be a strong predictor of HIV incidence in a recent meta-analysis [22]. The seemingly sub-optimal efficiency of HIV testing programs at subnational levels may suggest the failure of HIV programs in some settings to reach those who are at higher risk of HIV. However, risk may have been affected when people with HIV (PWH) undergo ART which ensures viral suppression, thus preventing transmission. Another potential reason why the level of HIV testing did not match the level of HIV risk is that in settings with high prevalence, people with known HIV infection (possibly under treatment) do not necessarily need to seek HIV testing. A further similar survey might be useful to assess recent HIV testing among the at-risk population only (i.e., excluding PWH under ART), however our data do not allow us to do so.

This study carries several limitations. First, the self-reported nature of HIV testing uptake may be subject to recall and social desirability biases resulting to underreporting. Second, differential accuracy in self-reporting between socioeconomic groups might have biased our results. Evidence in cancer screening suggest that overreporting of self-reported screening is common among marginalized groups such as racial minorities [28]. If this also applies to HIV testing, this may have led to an under-estimation of the pro-rich inequalities and over-estimation of the pro-poor inequalities. While self-reported lifetime HIV testing was found to be highly sensitive (96-99%) [29], testing in the past 12 months may be prone to telescoping bias that may have led to over-reporting [30]. Third, the wealth index can only measure relative wealth within a country. However, it can measure long-term SEP and has also been found to be stable especially in the Global South. Lastly, some available DHS surveys were conducted before 2014 and may not have captured more recent patterns of inequalities.

Despite these limitations, to our knowledge, this is the first study to provide a comprehensive context of the socioeconomic inequalities in HIV testing in SSA by quantifying and mapping them at different geographical levels on both absolute and relative scales and by assessing the spatial efficiency of HIV testing at different levels. This study revealed the importance of monitoring inequalities at different geographical scales. First, national estimates are often use for funding allocations by donors, prioritization of programs and comparison of inequality metrics. Second, province-level estimates are essential for program implementation, and for within-country funding allocations. Lastly, fine-level estimates allow us to visualize small-scale heterogeneities to precisely target communities in need.

## Conclusions

Our results show the need to monitor inequalities and assess the efficiency of HIV testing services in reaching those who are at-risk of HIV at smaller geographical scales, beyond national estimates that may mask disparities. By providing estimates of such inequalities at national-, province-, and PSU-level, and by localizing their hotspots, these findings may help policymakers, local and international organizations to prioritize areas and groups that need HIV testing efforts, while increasing efficiency.

## Data Availability

Raw data from the DHS surveys used in this study are publicly available for academic research. Formatted and processed data supporting the findings of this study are available from the corresponding author on request.

https://www.dhsprogram.com/

## Authors’ contributions

P.A.A.-T., L.T. and K.J. conceived and discussed the study with inputs from G.C.-E. and T.B. P.A.A.-T. collated and processed the DHS. P.A.A.-T. conducted the analysis with inputs from G.C.-E., T.B., L.T. and K.J. P.A.A.-T. produced output figures and tables with inputs from G.C.-E, T.B., L.T. and K.J. All authors contributed to the interpretation of the results. P.A.A.-T. wrote the initial draft with inputs from L.T. and K.J. All authors contributed to subsequent revisions. All authors read and approved the final version of the manuscript.

## Disclaimer

Funding agency had no role in the study design, data collection and analysis.

## Additional files

Additional file 1: **Supplementary material**

### Description

**Table S1**. Total number of participants in Primary Sampling Units that have at least 10 samples.

**Figure S1 (A)**. PSU distribution among women A) sample size and B) proportion of self-reported uptake of recent (< 12 months) HIV testing.

**Figure S1 (B)**. PSU distribution among men A) sample size and B) proportion of self-reported uptake of recent (< 12 months) HIV testing.

**Figure S2 (A)**. Global Moran’s I statistic test by number of nearest neighbor (1-10) by inequality scales among women.

**Figure S2 (B)**. Global Moran’s I statistic test by number of nearest neighbor (1-10) by gender and inequality scales.

**Figure S3**. Correlation (Pearson’s correlation coefficient R and p-value) between weighted HIV prevalence and weighted self-reported recent (< 12 months) uptake of HIV testing at the national level in 25 sub-Saharan African countries by gender.

**Figure S4**. Correlation (Pearson’s correlation coefficient R and p-value) between weighted HIV prevalence and weighted self-reported recent (< 12 months) uptake of HIV testing at the regional level in 25 sub-Saharan African countries by gender.

**Figure S5**. Sensitivity analysis of local spatial autocorrelation of socioeconomic inequalities in self-reported (<12 months) uptake of HIV testing as Local Getis-Ord Gi* at Primary Sampling Unit (PSU) level (2 nearest neighbors) with at least 20 participants across sub-Saharan African countries.

**Figure S6**. Sensitivity analysis of local spatial autocorrelation of socioeconomic inequalities in self-reported (<12 months) uptake of HIV testing as Local Getis-Ord Gi* at Primary Sampling Unit (PSU) level (2 nearest neighbors) with at least 20 participants across sub-Saharan African countries.

**Figure S7**. Sensitivity analysis of local spatial autocorrelation of HIV testing socioeconomic inequalities as Local Getis-Ord Gi* at PSU level (two nearest neighbors) across sub-Saharan African countries with surveys between 2011 and 2014.

**Figure S8**. Sensitivity analysis of local spatial autocorrelation of HIV testing socioeconomic inequalities as Local Getis-Ord Gi* at PSU level (two nearest neighbors) across sub-Saharan African countries with surveys between 2015 and 2019.

