## Appendices for "Investigating inequalities in HIV testing in sub-Saharan Africa: insights from a spatial analysis of 25 countries"

**Table S1. Total number of participants in Primary Sampling Units that have at least 10 samples.**

| ISO | Country | Year | Female (n) | Male (n) |
| --- | --- | --- | --- | --- |
| AO | **Angola**  (WCA) | **2015-16** | 11736 | 2489 |
| BI | **Burundi**  (ESA) | **2016-17** | 16046 | 5439 |
| CD | **Congo DR**  (WCA) | **2013-14** | 9110 | 3372 |
| CI | **Côte d’Ivoire**  (WCA) | **2011-12** | 1781 | 1402 |
| CM | **Cameroon**  (WCA) | **2018** | 14393 | 6080 |
| ET | **Ethiopia**  (ESA) | **2019** | 9711 | 7895 |
| GA | **Gabon**  (WCA) | **2012** | 7422 | 4668 |
| GH | **Ghana**  (WCA) | **2014** | 8223 | 1404 |
| GN | **Guinea**  (WCA) | **2018** | 7383 | 1164 |
| LB | **Liberia**  (WCA) | **2019** | 8897 | 2669 |
| LS | **Lesotho**  (ESA) | **2014** | 6370 | 907 |
| ML | **Mali**  (WCA) | **2018** | 5541 | 1106 |
| MW | **Malawi**  (ESA) | **2015-16** | 24204 | 3549 |
| MZ | **Mozambique**  (ESA) | **2015** | 7113 | 4163 |
| NM | **Namibia**  (ESA) | **2013** | 9160 | 1760 |
| RW | **Rwanda**  (ESA) | **2014-15** | 12440 | 4808 |
| SL | **Sierra Leone**  (WCA) | **2019** | 14654 | 4239 |
| SN | **Senegal**  (WCA) | **2017** | 15952 | 3796 |
| TD | **Chad**  (WCA) | **2014-15** | 4389 | 607 |
| TG | **Togo**  (WCA) | **2013-14** | 8926 | 2802 |
| TZ | **Tanzania**  (ESA) | **2011-12** | 10037 | 6754 |
| UG | **Uganda**  (ESA) | **2011** | 9287 | 7707 |
| ZA | **South Africa**  (ESA) | **2016** | 6581 | 634 |
| ZM | **Zambia**  (ESA) | **2018** | 13276 | 11740 |
| ZW | **Zimbabwe**  (ESA) | **2015** | 9876 | 8259 |

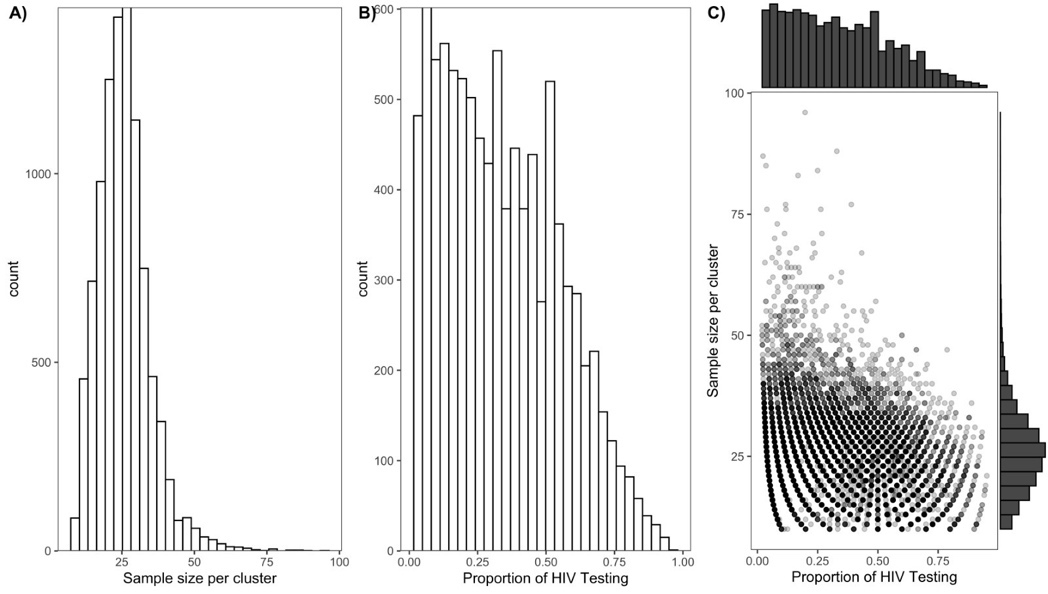

**Figure S1 (A). PSU distribution among women A) sample size and B) proportion of self-reported uptake of recent (< 12 months) HIV testing.**

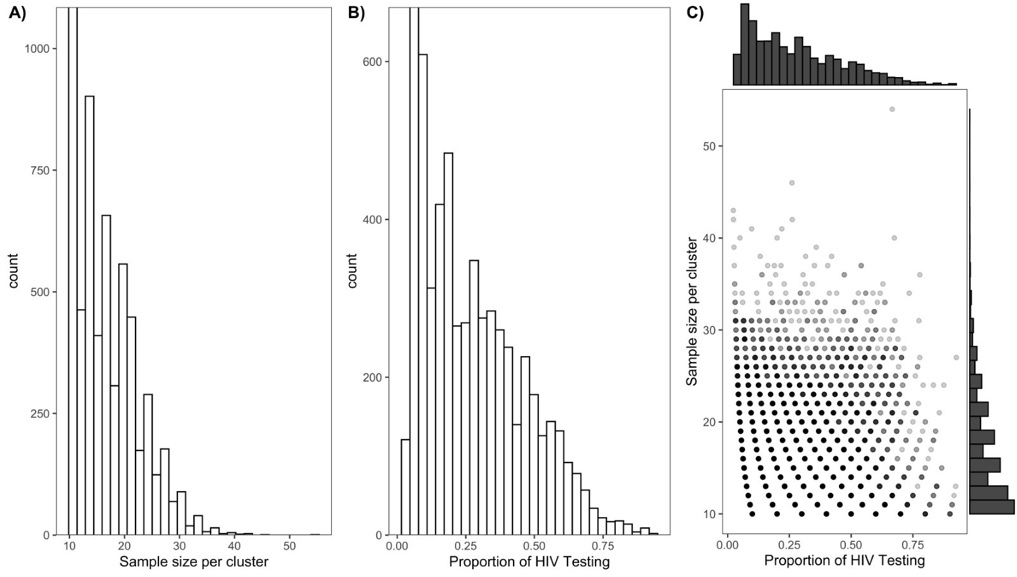

**Figure S1 (B). PSU distribution among men A) sample size and B) proportion of self-reported uptake of recent (< 12 months) HIV testing.**

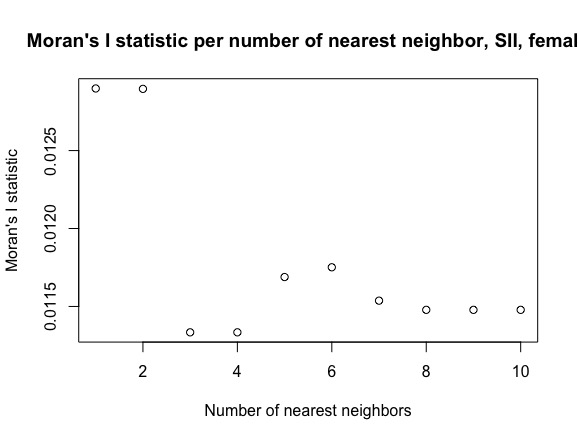

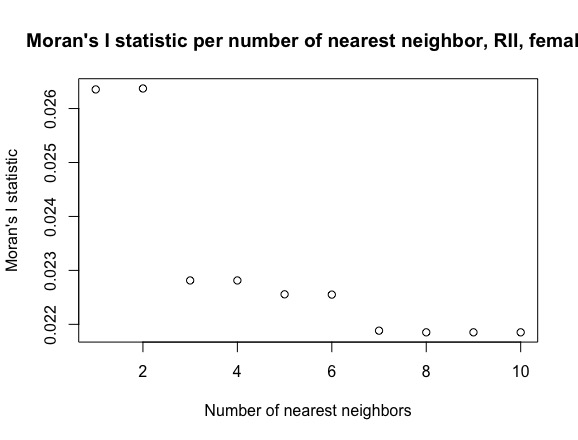

**Figure S2 (A). Global Moran’s I statistic test by number of nearest neighbor (1-10) inequality scales among women.**

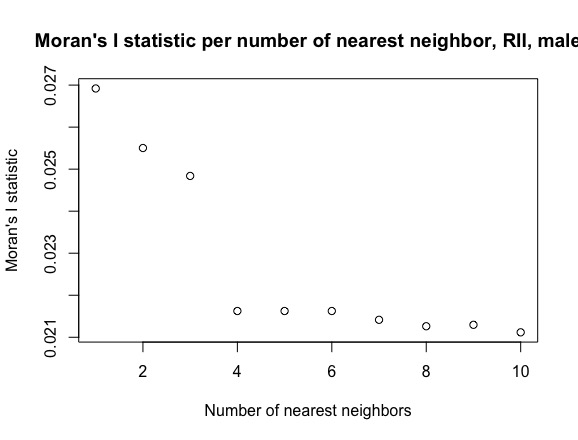

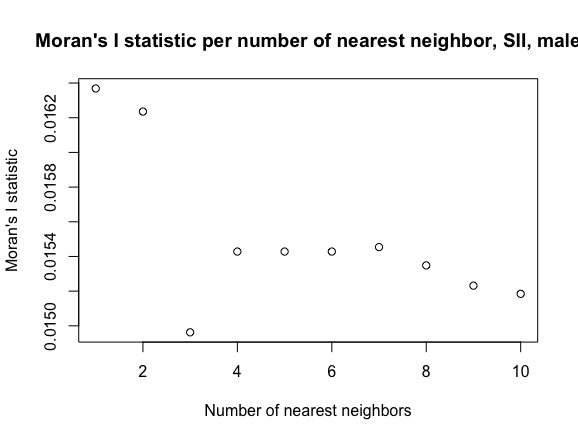

**Figure S2 (B). Global Moran’s I statistic test by number of nearest neighbor (1-10) by inequality scales among men.**

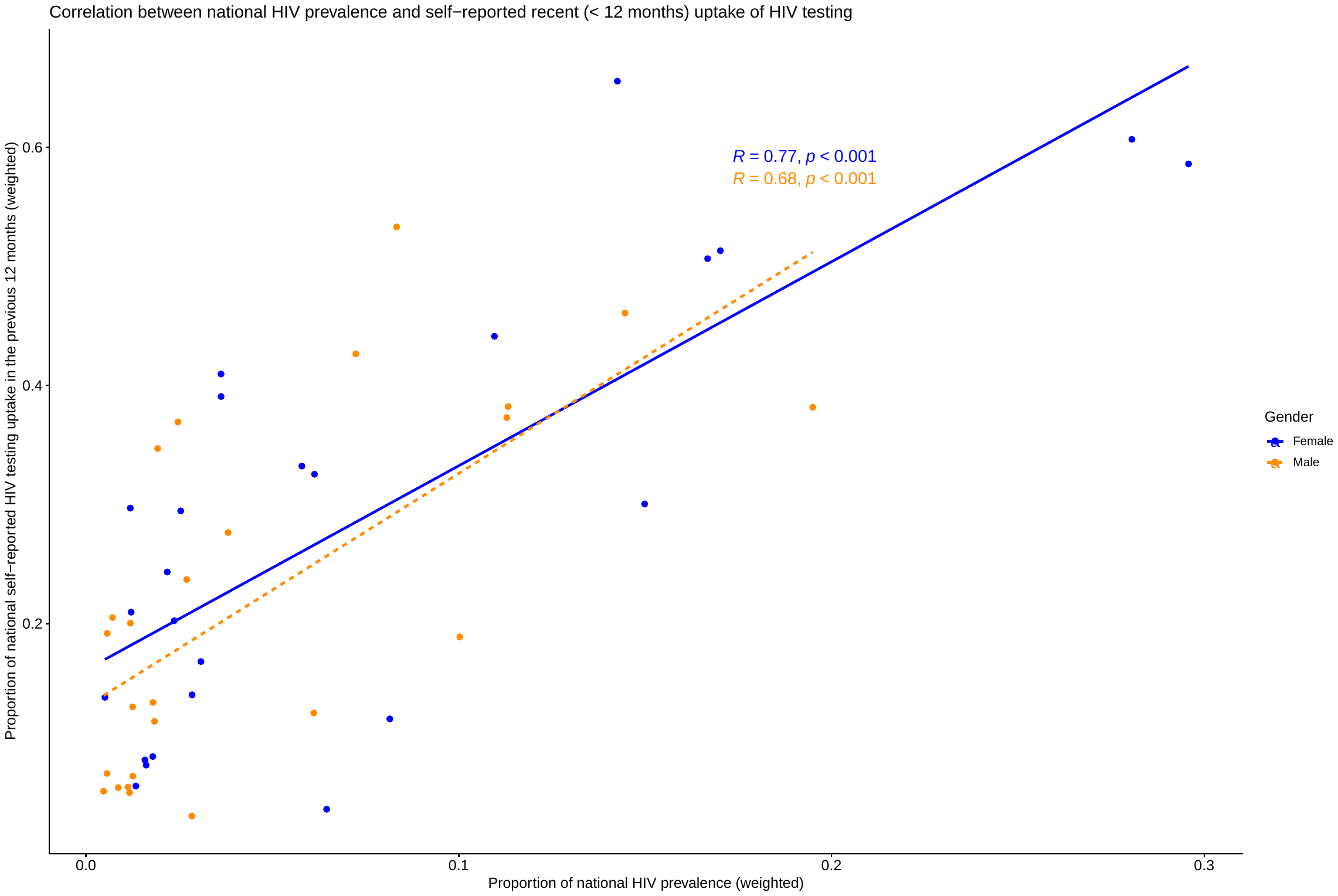

**Figure S3. Correlation (Pearson’s correlation coefficient R and p-value) between weighted HIV prevalence and weighted self-reported recent (< 12 months) uptake of HIV testing at the national level in 25 sub-Saharan African countries by gender.**

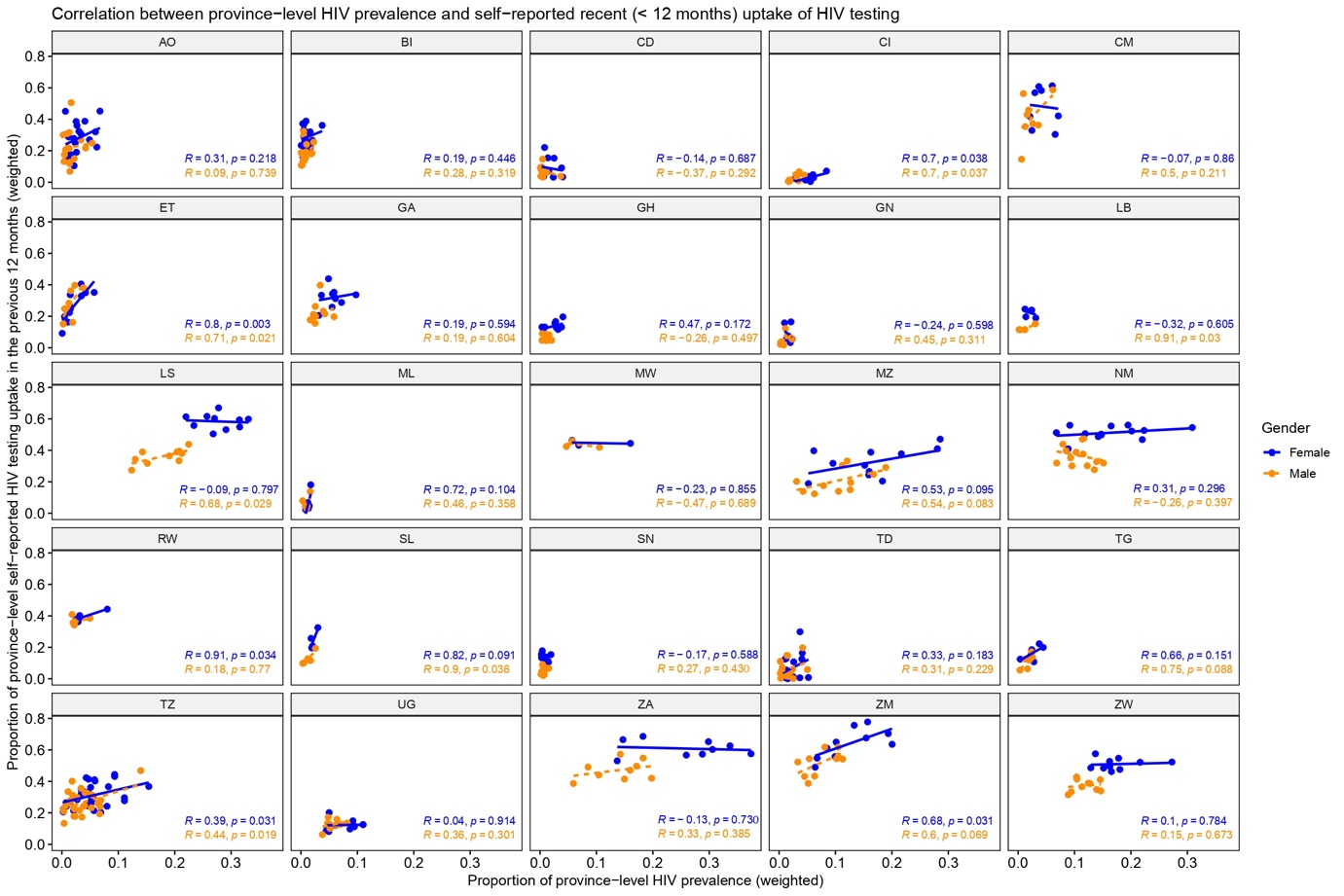

**Figure S4. Correlation (Pearson’s correlation coefficient R and p-value) between weighted HIV prevalence and weighted self-reported recent (< 12 months) uptake of HIV testing at the regional level in 25 sub-Saharan African countries by gender.** Only included regions with both the HIV biomarker and HIV testing variables.

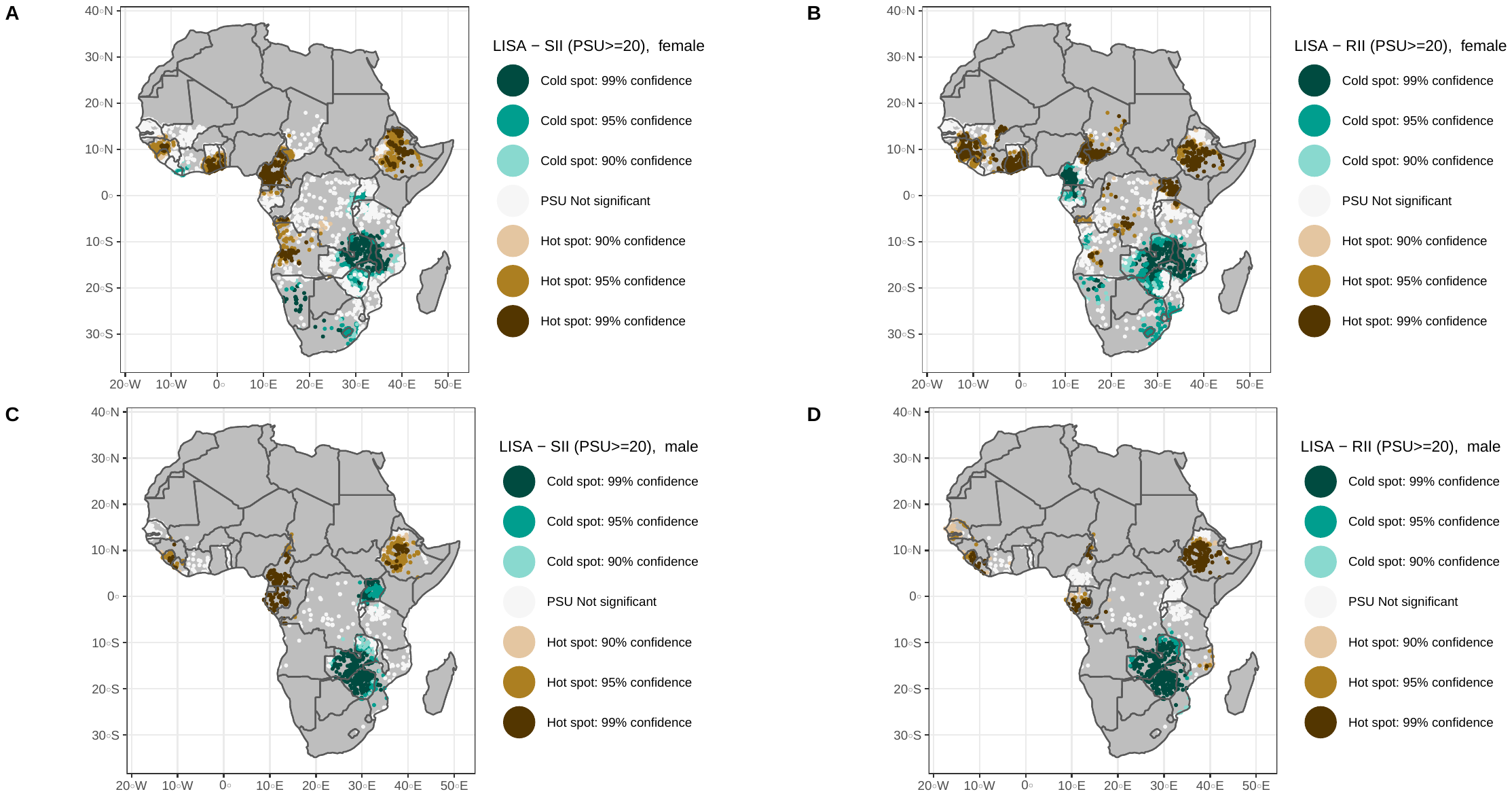

**Figure S5. Sensitivity analysis of local spatial autocorrelation of socioeconomic inequalities in self-reported (<12 months) uptake of HIV testing as Local Getis-Ord Gi* at Primary Sampling Unit (PSU) level (2 nearest neighbors) with at least 20 participants across sub-Saharan African countries.** Spatial clustering at the A) and C) absolute scales and B) and D) relative scales among women and men, respectively. Only PSUs with a sample size of at least 20 and more than one wealth quintile were included.

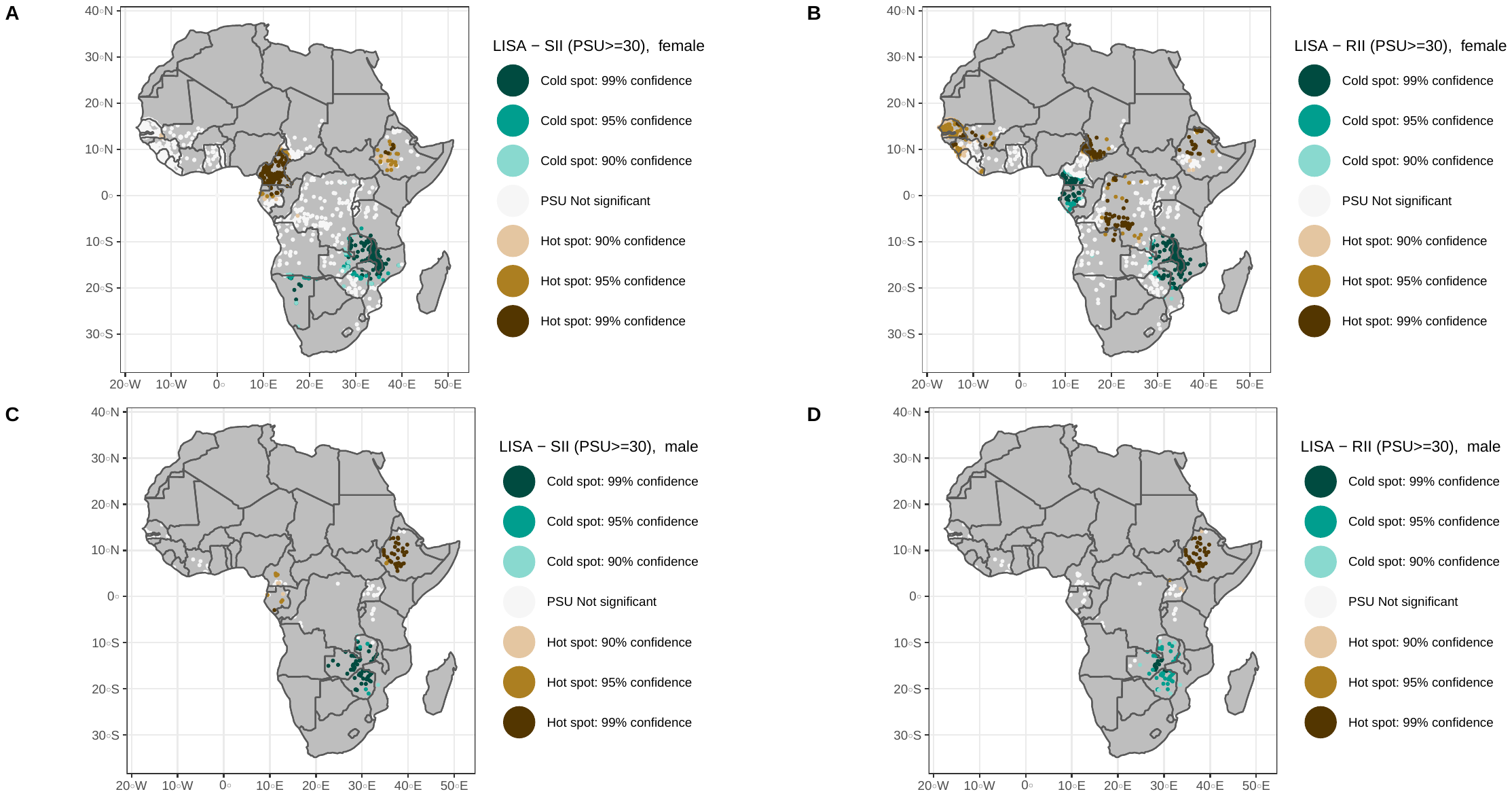

**Figure S6. Sensitivity analysis of local spatial autocorrelation of socioeconomic inequalities in self-reported (<12 months) uptake of HIV testing as Local Getis-Ord Gi* at Primary Sampling Unit (PSU) level (2 nearest neighbors) with at least 30 participants** **across sub-Saharan African countries.** Spatial clustering at the A) and C) absolute scales and B) and D) relative scales among women and men, respectively. Only PSUs with a sample size of at least 30 and more than one wealth quintile were included.

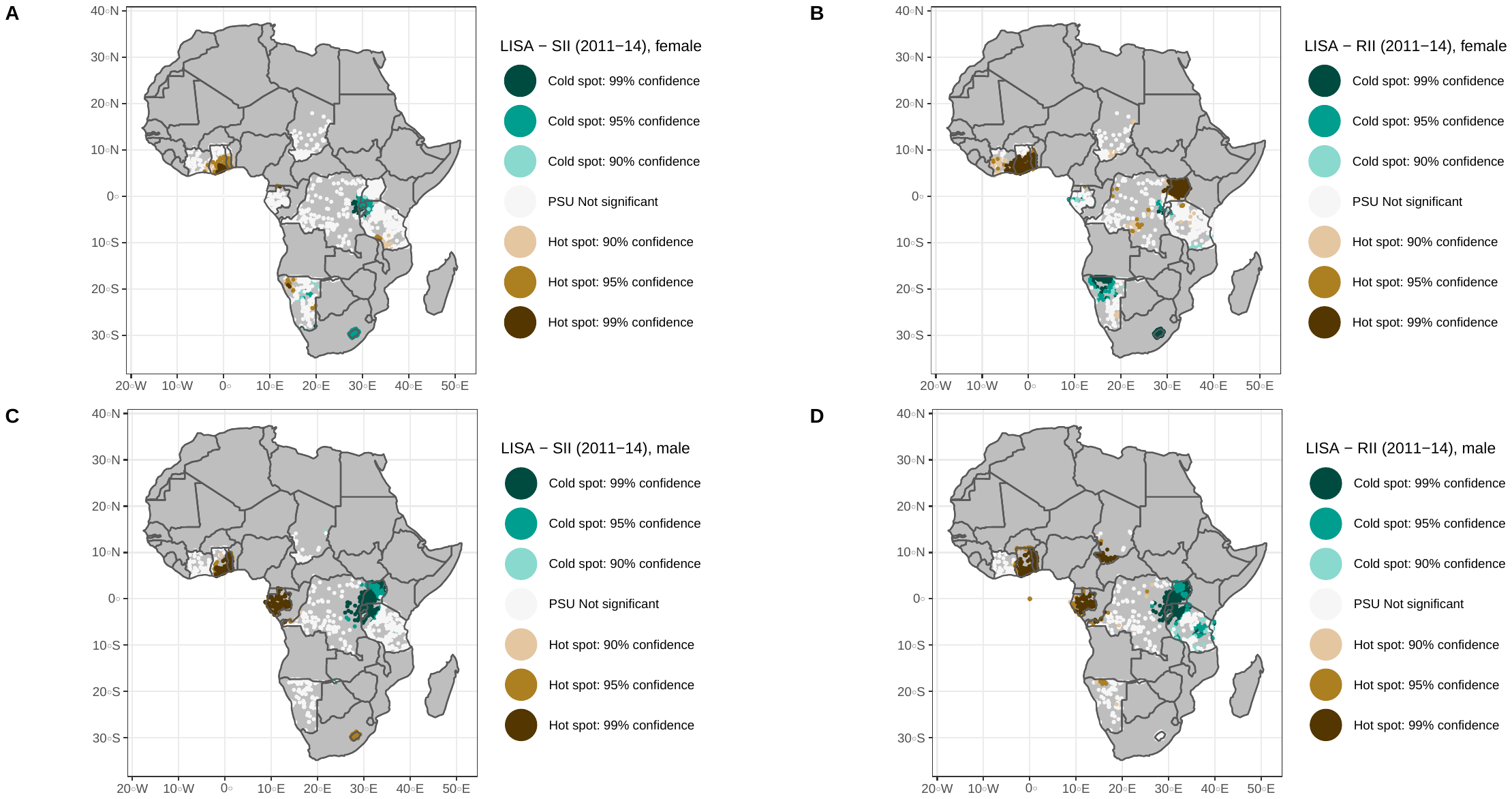

**Figure S7.** **Sensitivity analysis of local spatial autocorrelation of HIV testing socioeconomic inequalities as Local Getis-Ord Gi* at PSU level (two nearest neighbors) across sub-Saharan African countries with surveys between 2011 and 2014.** Spatial clustering at the A) and C) absolute scales and B) and D) relative scales among women and men, respectively. Only PSUs with a sample size of at least 10 and more than one wealth quintile were included.

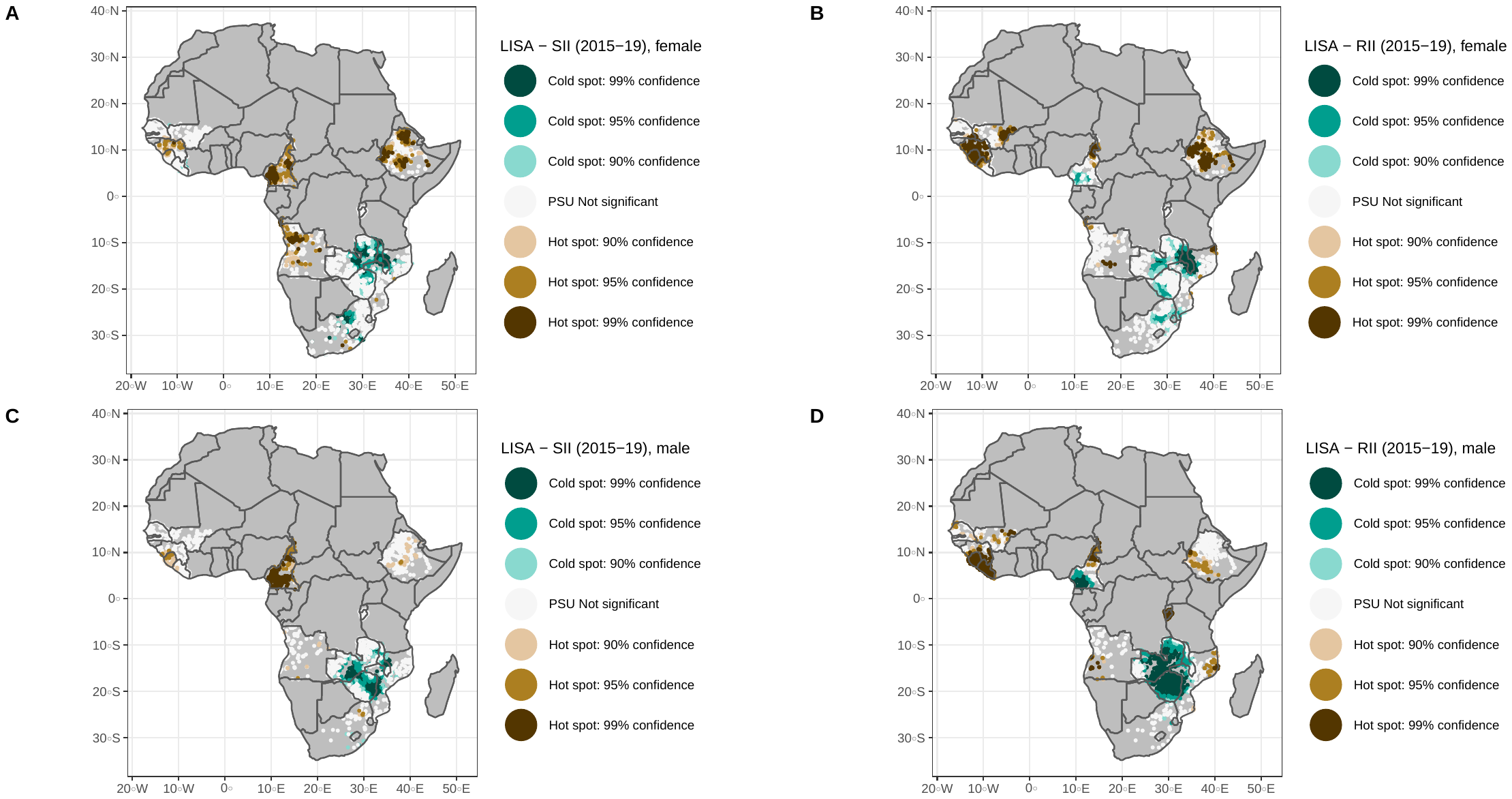

**Figure S8.** **Sensitivity analysis of local spatial autocorrelation of HIV testing socioeconomic inequalities as Local Getis-Ord Gi* at PSU level (two nearest neighbors) across sub-Saharan African countries with surveys between 2015 and 2019.** Spatial clustering at the A) and C) absolute scales and B) and D) relative scales among women and men, respectively. Only PSUs with a sample size of at least 10 and more than one wealth quintile were included.
